# SARS-CoV-2 variants: levels of neutralisation required for protective immunity

**DOI:** 10.1101/2021.08.11.21261876

**Authors:** Deborah Cromer, Megan Steain, Arnold Reynaldi, Timothy E Schlub, Adam K Wheatley, Jennifer A Juno, Stephen J Kent, James A Triccas, David S Khoury, Miles P Davenport

## Abstract

A number of SARS-CoV-2 variants of concern (VOC) have been identified that partially escape serum neutralisation activity elicited by current vaccines. Recent studies have also shown that vaccines demonstrate reduced protection against symptomatic infection with SARS-CoV-2 variants. Here we integrate published data on in vitro neutralisation and clinical protection to understand and predict vaccine efficacy against existing SARS-CoV-2 variants. We find that neutralising activity against the ancestral SARS-CoV-2 is highly predictive of neutralisation of the VOC, with all vaccines showing a similar drop in neutralisation to the variants. Neutralisation levels remain strongly correlated with protection from infection with SARS-CoV-2 VOC (r=0.81, p=0.0005). We apply an existing model relating in vitro neutralisation to protection (parameterised on data from ancestral virus infection) and find this remains predictive of vaccine efficacy against VOC once drops in neutralisation to the VOC are taken into account. Modelling of predicted vaccine efficacy against variants over time suggests that protection against symptomatic infection may drop below 50% within the first year after vaccination for some current vaccines. Boosting of previously infected individuals with existing vaccines (which target ancestral virus) has been shown to significantly increase neutralising antibodies. Our modelling suggests that booster vaccination should enable high levels of immunity that prevent severe infection outcomes with the current SARS-CoV-2 VOC, at least in the medium term.

## Introduction

The global spread of SARS-CoV-2 has resulted in significant morbidity, mortality, and social disruption. Several vaccines have been deployed that protect against symptomatic SARS-CoV-2 infection (reviewed in^1^). Vaccines in current use incorporate the ancestral (Wuhan-like) virus or viral spike protein as an immunogen. Both vaccination and prior infection have been shown to provide a degree of protection against symptomatic and severe infection with essentially homologous virus^2-4^. Recently, several SARS-CoV-2 variants of concern (VOC) have emerged that display increased transmissibility and / or reduced in vitro neutralisation by sera from subjects infected with the ancestral strain or immunised with current vaccines^5-7^. Initial reports from clinical trials or from breakthrough community infections suggest that current vaccines may be less protective against symptomatic infection with some SARS-CoV-2 variants^8-12^. In addition, studies also demonstrate that waning antibody levels correlate with reduced protection over time^13,14^. Thus, a major question is the extent to which existing vaccines are likely to protect against variants of concern and how existing vaccines might be used to combat the threat of variants.

Current vaccines have been shown to elicit different levels of neutralising antibody in vaccinated subjects, ranging from ∼0.2-fold to ∼4-fold of the levels seen in early convalescence^15-17^. Studies analysing vaccine-induced neutralising antibody responses have reported varying levels of reduction in neutralisation titre against variants of concern (VOC). However, the lack of a standardised assay to measure in vitro neutralisation means that the absolute serum dilution titres sufficient to neutralise either ancestral or variant viruses differ considerably between laboratories^18^. There are also now a number of studies reporting vaccine efficacy against variants, which also indicate a variable reduction in efficacy^10,19^. Our previous work has shown a correlation between neutralising antibody levels and protection from SARS-CoV-2 infection and has derived a model for predicting vaccine efficacy from mean neutralisation titres^14^. However, this model was developed based on neutralisation and protection from the ancestral virus, and whether this remains predictive for efficacy against SARS-CoV-2 VOC has not been determined. A predictive model of vaccine protection against SARS-CoV-2 variants would allow us to forecast the ongoing utility of current vaccines in the face of emerging variants, and to model the utility of boosters and other strategies to extend the duration of vaccine protection.

In this study we aim to determine the utility of existing vaccines (targeting ancestral virus) for protecting against VOC. We first compare the observed in vitro neutralisation titres from convalescent subjects and vaccinees immunised with 4 different vaccines against all current SARS-CoV-2 VOCs: namely alpha (B.1.1.7), beta (B.1.351), gamma (P.1) and delta (B.1.617.2). We find that the reported reductions in serum neutralisation activity against SARS-CoV-2 VOC in vitro is independent of whether immunity was established by prior infection or vaccination, and was consistent between vaccine platforms. Combining this in vitro data with clinical studies of vaccine efficacy, we show that in vitro neutralisation is significantly correlated with protection from infection with SARS-CoV-2 variant virus in vaccinated individuals. This relationship is consistent with the existing model relating in vitro neutralisation titre to observed protection, suggesting this model can be used to predict vaccine efficacy against SARS-CoV-2 variants. Using this model, we study the effects of immune boosting of previously infected subjects. This work suggests that losses of vaccine effectiveness in the face of evolving variant viruses and waning immunity may be partially offset by timely booster immunisations.

## Results

### Cross reactivity of neutralising antibodies against SARS-CoV-2 variants of concern

It has previously been shown that neutralising antibodies against ancestral virus correlate with vaccine efficacy^14,20-22^. In order to determine if neutralisation responses continue to correlate with protection for VOC, a robust estimate of the loss of neutralising activity against VOC for each vaccine is needed. We examined the loss of neutralisation against VOC by combining data from 16 published studies which directly compared neutralisation titre against ancestral (Wuhan-like / D614G) strains and the VOC (see supplementary table 1)^5,7,23-36^. Different laboratories used distinct in vitro assays to measure neutralisation of SARS-CoV-2 (supplementary table 1)^37^, and reported considerably different means and fold-changes in neutralisation to variants (Fig 1a). However, within a given study the observed drop in neutralisation titre for a given variant was very similar for both convalescent and vaccinee serum, suggesting a strong study-specific (or assay-specific) effect (supplementary Figure S1). To explore the contribution of assay-specific and vaccine-specific effects, we used a regression model (with censoring) to aggregate data from all studies and included potential for assay-specific, variant-specific and vaccine-specific effects (supplementary analysis and methods). From this regression we found that neutralisation against ancestral virus was strongly associated with neutralisation against a particular variant, and that after adjusting for variant and laboratory effects, whether immunity was acquired through infection or vaccination (and which vaccine was used) was not a significant factor associated with the loss of neutralisation (p=0.26, likelihood ratio test) (Figure 1b and Supplementary Figure 2). Importantly, this does not imply that all vaccinee serum neutralised variants equally well, rather that the loss of neutralisation of a variant (i.e. the average fold drop in neutralisation against a given variant compared to ancestral virus) did not differ between vaccines. Since all vaccines suffer a similar loss of variant recognition, this demonstrates that neutralisation against the ancestral virus can be used to predict neutralisation against the current variants of concern (Figure 1b, Supplementary Figure S2). The average drop in neutralisation level for different variants is shown in Supplementary Figure S3.

**Figure 1:**
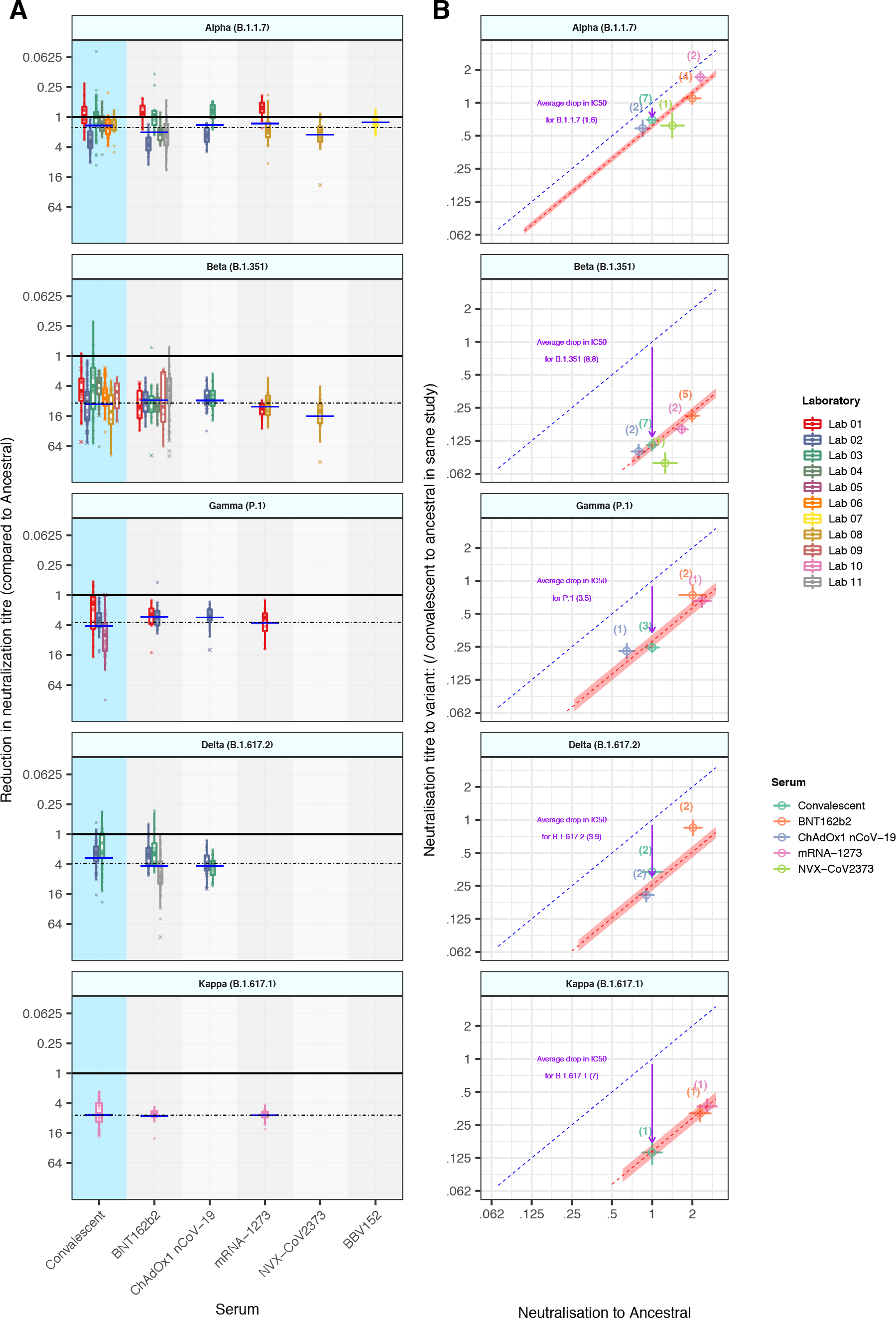
In vitro neutralisation of SARS-CoV-2 variants: (a) The change in neutralisation titre between the ancestral virus and different SARS-CoV-2 variants for either convalescent individuals (left) or those immunised with different vaccines is shown. Individual colours reflect different studies / laboratories (described in detail in Supplementary Table 1). Solid dots indicate where titres were measurable for both ancestral and variant neutralisation. Crosses indicate where one titre fell below the limit of detection for that assay. Different studies estimate quite different changes in neutralisation titre even for the same vaccine / variant combination. The dashed horizonal line indicates the weighted mean drop in titre for a given variant (across all vaccine and convalescent samples), and horizontal bars indicate the weighted mean titre for a given vaccine / variant combination. (b) The mean neutralisation titre against the ancestral virus (x-axis) is highly correlated with mean neutralisation titre against the VOC (y-axis). The predicted line for a 1:1 relationship is indicated (dashed blue line). The observed mean drop in neutralisation titre across all vaccines and convalescent subjects is indicated by an arrow, and the predicted levels of variant neutralisation are indicated by a dashed red line (shading indicates 95% CI) are shown. The points indicated are the mean neutralisation levels for a given vaccine / variant combination, averaging across available studies (number of studies indicated).

### Predicting vaccine efficacy against SARS-CoV-2 variants

Several studies have now shown reduced efficacy of vaccination against infection with SARS-CoV-2 variants^10,11,19,38-41^ (supplementary table 2). These studies incorporated a variety of study designs, including both randomised controlled trials (RCT)^10,19,40,41^ and observational case-control studies^38,39^. Vaccine efficacy against the variants was assessed either by separately analysing the number of infections with variant virus^38^ or on the assumption (based on epidemiological data) that the vast majority of infections were with variant virus^19,40^. In addition to these differences in study design, other factors such as the definitions of mild, moderate, and severe infection also differed by study (Supplementary Table 2). We used the correlation between ancestral and variant neutralisation titres described above (Figure 1b) to estimate the neutralisation level of each vaccine-variant combination, and related this to the protection observed for that vaccine-variant combination (Figure S4). Despite the variability in study design, we find that predicted serological neutralisation activity against each variant elicited by vaccines is significantly correlated with protection from COVID-19 (r=0.81, p=0.0005, Spearman, Supplementary Figure S4).

To test whether our previously developed predictive model could also be used to predict efficacy against VOCs, we took the same model (parameterised from ancestral virus) and applied the mean drop in neutralisation titres estimated above (Supplementary Figure 3) to effectively shift the curve for each variant (see Supplementary Methods). Figure 2 plots the neutralisation titre for each vaccine against the ancestral virus (x-axis) and the predicted efficacy against each VOC (y-axis). The predicted efficacy (solid line) and 95% confidence intervals (shaded) are shown, along with the reported vaccine efficacies against symptomatic infection with each VOC as reported in clinical studies (figure 2a, Supplementary Table 2). These efficacy studies show very good agreement with the model predictions for each vaccine, with 13/14 efficacy studies falling within the 95% confidence interval of our model. Importantly, these confidence intervals reflect the known sources of uncertainty, including (i) uncertainty in estimates of neutralisation against ancestral virus, (ii) uncertainty in estimates of the drop in neutralisation for each variant and (iii) model-related uncertainty (Supplementary Table 3). This means that we can use a single estimate of the mean neutralisation level of a vaccine against ancestral virus (as reported in the phase I/II trial) to provide both a point estimate of the efficacy of the same vaccine against the VOCs, as well as confidence limits on this point estimate. This model provides a clear prediction (and lower bound) of vaccine efficacy against the VOC for a given neutralisation level against ancestral virus.

**Figure 2:**
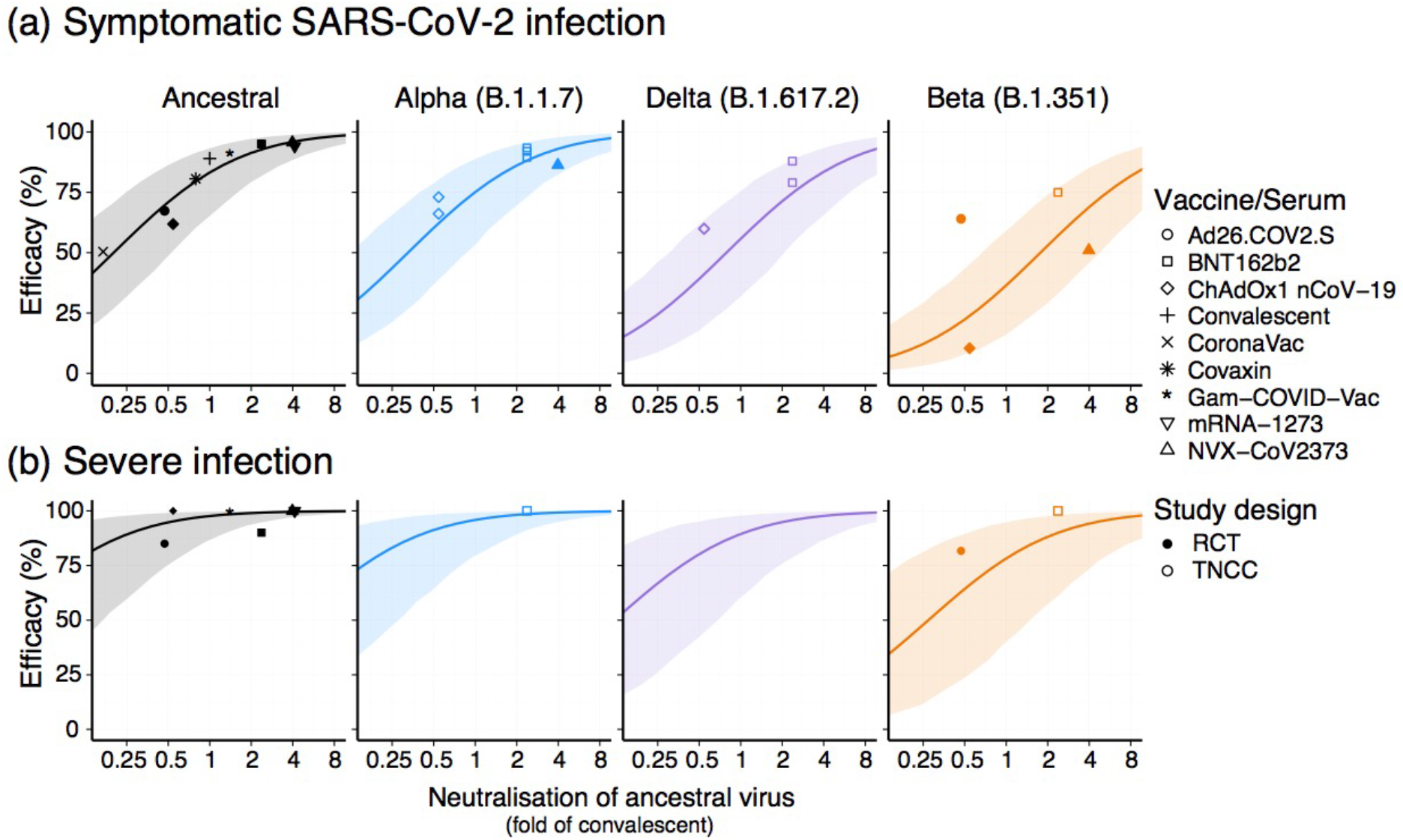
Predicting vaccine efficacy against SARS-CoV-2 variants: The relationship between mean neutralisation level to ancestral SARS-CoV-2 and protection against (a) symptomatic and (b) severe infection with different variants is shown. The line indicates the model prediction of efficacy for a given level of neutralisation against ancestral virus. Shading indicates 95% confidence interval based on uncertainties in measuring mean neutralisation titre against ancestral virus, the loss of neutralisation against each variant and in model parameters. Individual points shown represent results of different studies of vaccine efficacy against ancestral virus (black) or SARS-CoV-2 variants (different study designs are shown as solid (RCT) or open (TNCC) shapes). Details of studies of ancestral virus are outlined in reference^14^ (all of which are RCT) and for VOC this is outlined in supplementary table 2.

Predicting vaccine efficacy against severe SARS-CoV-2 infection is significantly more challenging, due to the low numbers of severe infections captured in most efficacy studies^14^. However, even with this limitation, vaccination has been shown to provide significantly better protection against severe disease than against symptomatic SARS-CoV-2 infection^42^. Thus we next considered our model’s prediction of efficacy against severe outcomes with VOC (Figure 2b). The 95% confidence intervals are substantially broader for severe than for symptomatic SARS-CoV-2 infection, indicating the greater uncertainty in the data on protection from severe disease. Further, the point estimates of efficacy from the clinical studies contain considerable uncertainty (Supplementary Table 4). Even so, all but one of the efficacy studies falls within or above the predicted efficacy confidence limits. It should be noted that our model assumes neutralisation alone drives protection against severe disease, but it is likely that other cellular responses play a critical role in modulating disease severity, and thus the model may underestimate efficacy against severe COVID-19.

### Boosting existing SARS-CoV-2 antibody responses

The models above and emerging data on breakthrough infections suggest that booster vaccinations may eventually be required to both augment waning immunity and boost responses to variants. A major question is whether boosting with existing vaccines (that all currently incorporate only the ancestral spike) will be effective in providing protection against the variants. A number of studies have compared neutralising antibody responses after vaccination of previously infected individuals versus naïve individuals^43-47^. These have found that vaccination with a single dose of mRNA vaccine is sufficient to boost responses in previously infected individuals (and indeed a second dose has minimal effect)^45,46^. For example, the Phase I/II studies of BNT162b2 or mRNA-1273 show that vaccination of naïve individuals with the standard two dose regimen leads to a mean neutralisation titre of approximately 2 and 4-fold that of convalescent plasma, respectively. However, vaccination of convalescent individuals led to between 2 and 10-fold higher neutralisation levels than that seen in vaccinated naïve individuals (Figure 3, Supplementary Table S4)^43-48^ and additionally, improved the level of cross-reactivity to variants^44-47^. Although the level of increase varied between studies, vaccination of convalescent individuals led to boosting of neutralisation levels to approximately 12-fold (range, 6.1-fold to 29-fold) higher than that seen in early convalescence (and higher than that seen in any current vaccination regimen)(Figure 3).

**Figure 3:**
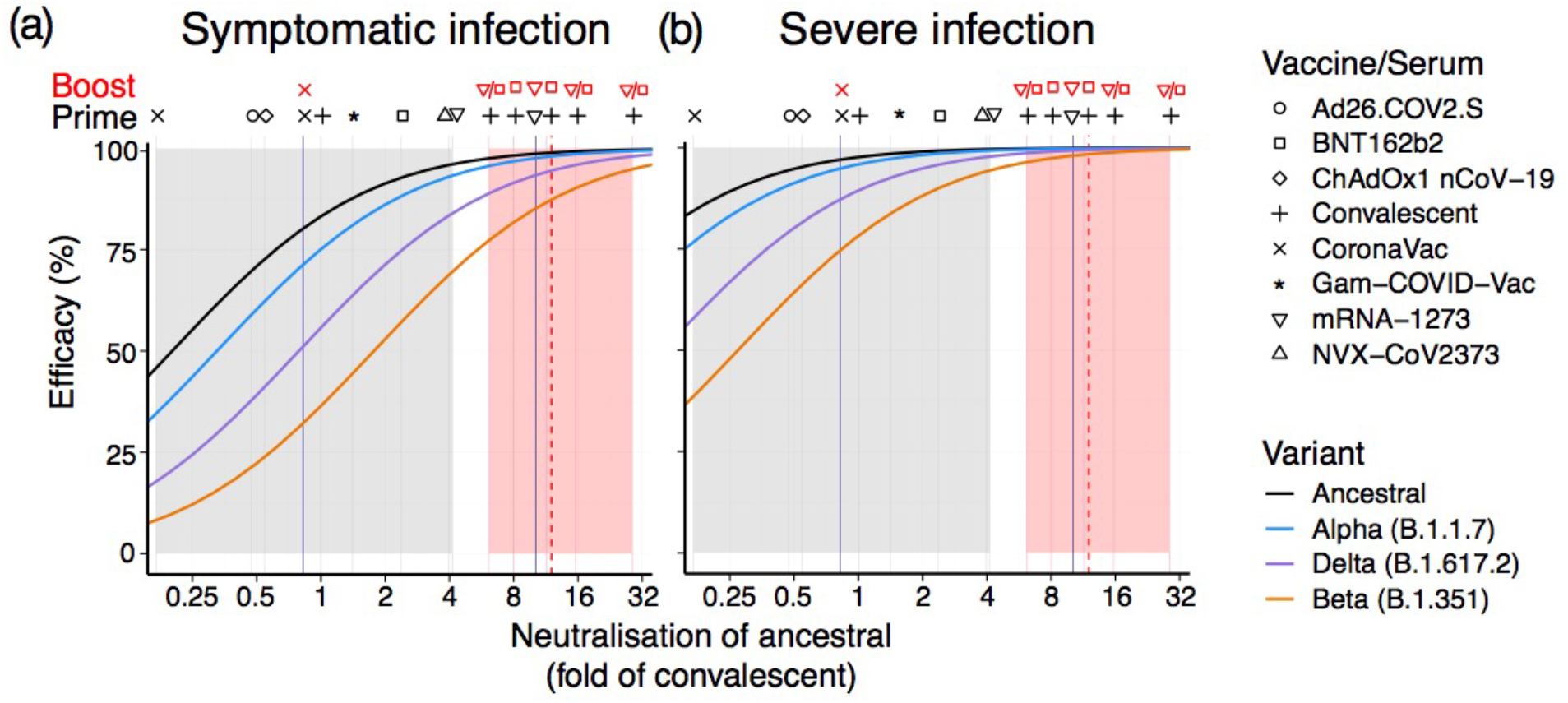
Predicted impact of boosting neutralising antibody responses: The observed levels of neutralising antibodies against ancestral virus following initial vaccination (grey) as well as the effects of boosting of previously infected individuals (red) are shown. Results for individual studies are indicated as vertical lines, and symbols above the lines indicate the vaccine(s) used and infection history. The geometric mean level of boosting seen in previously infected individuals (from all studies) is shown as a dashed red line. Shaded areas indicate the range of mean neutralisation levels observed following vaccination of naïve (grey) or previously infected (red) individuals. Two studies of boosting of previously vaccinated individuals are shown as vertical blue lines (vaccines indicated by symbols above). The modelled relationship between neutralisation and protection from ancestral (black) or VOC are shown as coloured sloped lines for either (A) any symptomatic SARS-CoV-2 infection or (B) severe infection.

We use our existing model to predict the impact of boosting, with existing vaccines targeting ancestral virus, on protection against symptomatic (figure 3a) and severe infection with SARS-CoV-2 VOC (figure 3b). This prediction assumes that the relationship between neutralisation and protection continues to hold after boosting and that the drop in titre against the variants is also similar after boosting^47^. As noted above, boosting may actually increase cross reactivity against VOC, and thus the protection shown here is likely a minimum bound on the true protection conferred from boosting. Regardless, this suggests that boosting of previously infected individuals using existing vaccines should lead to both higher neutralising titres and higher protection from symptomatic and severe COVID-19 (figure 3), consistent with a recent study of vaccine efficacy after vaccination of either naïve or previously infected individuals^49^.

There is currently more limited data available on the effects of boosting in vaccinated individuals (blue vertical lines, Figure 3). Wu et al^50^ showed that a third dose of mRNA-1273 delivered 6 months after the initial vaccination boosted neutralisation levels by around 23-fold compared to the pre-boost levels, or around 2.5-fold higher than vaccination of naïve individuals^48^. Pan et al^51^ have studied the effects of a third dose of CoronaVac delivered either 1 month or 6 months after the second dose. They found that a third vaccination at 6 months boosted responses approximately 3 to 5-fold higher than seen after the initial two dose regime. Interestingly, boosting at 1 month after the initial two dose regime led to only a 1.3-2.1-fold increase, suggesting that delayed boosting may be required. The level of boosting of vaccinated individuals is indicated as vertical blue lines in Figure 3.

Previous studies have shown a decrease in neutralising antibody titres over the first 8 months of infection, with a half-life of around 3-4 months^52^, and recent work has also shown a decrease in vaccine protection over this period^13^. This waning immunity, coupled with the drop in neutralisation titres to the VOC, has the potential to reduce protection over the first year after vaccination or infection (Figure 4)^13,14^. However, vaccination of previously infected individuals or boosting of previously vaccinated individuals has the potential to raise neutralising antibody levels above those seen after primary vaccination. Assuming that the decay of neutralisation titres after boosting is consistent with decay after primary infection^52-54^, vaccination of convalescent individuals is predicted to provide 69% protection from symptomatic infection (lower bound CI: 47%) and 94% protection from severe infection (lower bound CI: 74%) even against the most escaped VOC (beta) 6-months after boosting (figure 4)^49^. Although there are limited data on the effects of boosting of vaccinated individuals, preliminary data suggests that boosting with mRNA vaccines may be capable of achieving similar levels of neutralising responses (Figure 3)^50^. Further work is required to understand the optimal vaccination and boosting schedules that might achieve high levels of immunity to SARS-CoV-2 infection against all current variants for at least a 6 month interval (figure 4)^49^. Together this data and modelling suggest that vaccination of previously infected individuals, even with existing vaccines targeting ancestral virus, will provide robust protection against the current VOC, considerably prolonging the duration of efficacy of existing vaccines against these variants.

**Figure 4:**
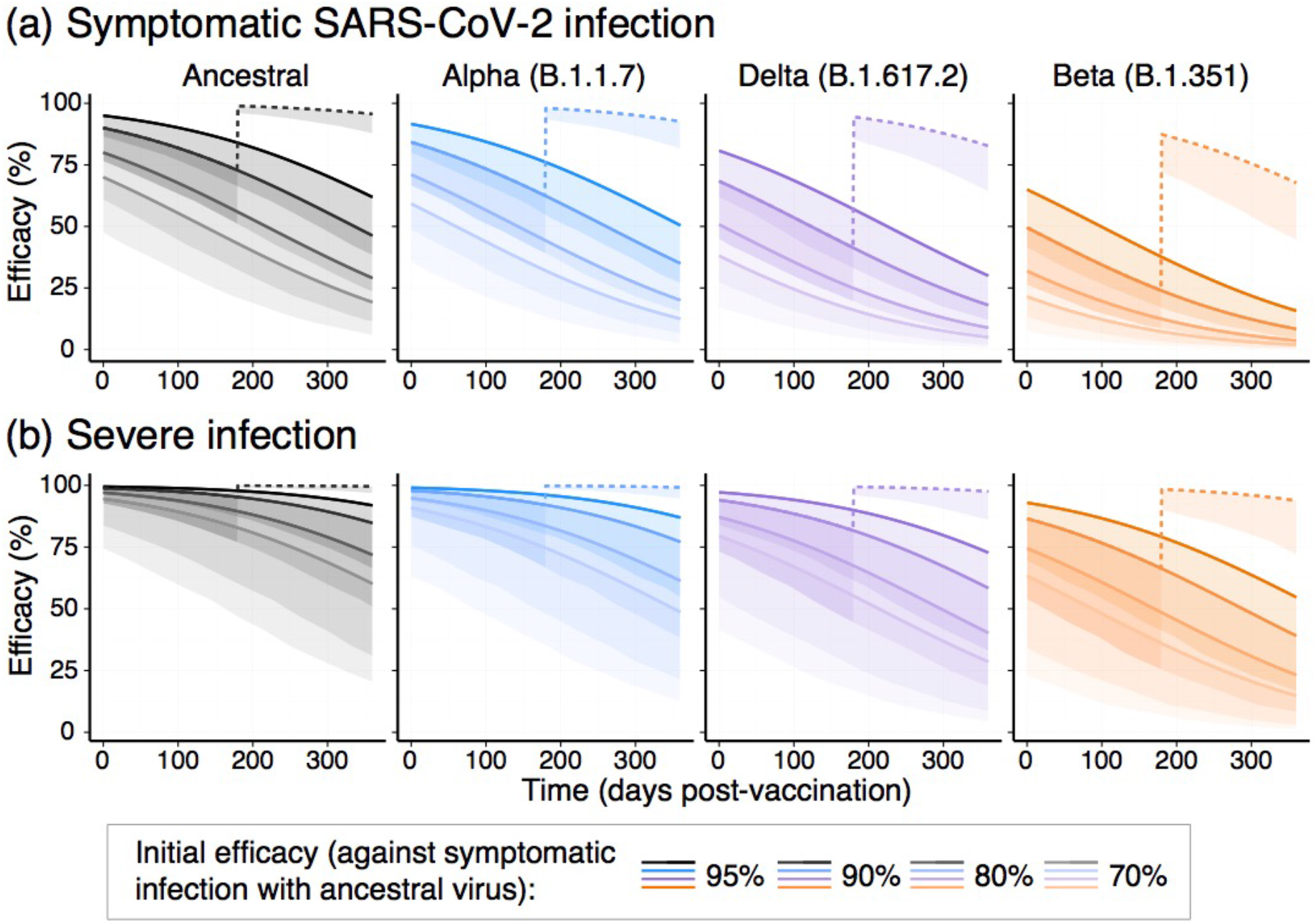
Predicted protection from SARS-CoV-2 infection and the impact of boosting: The predicted protection over time is shown for four hypothetical vaccines that initially provide 95%, 90%, 80% or 70% protection against symptomatic infection with the ancestral virus. It is assumed that neutralisation levels decay with a half-life of 108 days and variant neutralisation decreases as estimated above (see Supplementary figure S3). Solid lines are mean model prediction, and shading indicates lower bound of the 95% confidence interval (indicating the minimal predicted efficacy). Dashed line indicates the predicted impact of boosting previously infected individuals at six months after infection with BNT162b2 or mRNA-1273 (mean of all boosting studies, see Supplementary Table S4), and assumes decay after boosting is the same as after initial infection or primary vaccination.

## Discussion

The deployment of highly effective vaccines against SARS-CoV-2 is contributing to major reductions in disease in many countries. However, the spread of novel viral variants with increased transmissibility and potential for reduced susceptibility to neutralisation by vaccine-induced serological responses has raised concerns about the durability of vaccine protection. Thus, understanding how the emergence of SARS-CoV-2 variants affects the ability of vaccines to neutralise and protect against infection is an important priority. Here we integrate data from in-vitro neutralisation assays and efficacy studies incorporating a number of vaccines in widespread use. A major question is to understand whether immunity from existing vaccines shows a similar level of cross-reactivity to SARS-CoV-2 variants, or whether some vaccine platforms elicit more broadly cross-reactive neutralising antibody responses. After accounting for significant variation between the different assays used, we find that vaccine-elicited antibody responses display similar levels of cross reactivity between vaccine platforms, and show little difference to that seen in convalescent subjects. This is perhaps not surprising, since current vaccines incorporate very similar (ancestral) SARS-CoV-2 spike immunogens. It should be emphasised that the current analysis is limited by the lack of a standardised neutralisation assay or a direct comparison across a panel of serum from all existing vaccines, and as a result we cannot exclude minor variations in cross-reactivity.

Separate from *in-vitro* neutralisation, studies of *in-vivo* vaccine efficacy against SARS-CoV-2 variants are urgently needed to understand and predict future protection from infection. Studies to date have shown variable reductions in vaccine efficacy against circulating SARS-CoV-2 variants ^10,11,19,38-41^(Figure 2 and Supplementary Table 2). We previously derived a model to predict vaccine efficacy from in vitro neutralisation titre against (ancestral) SARS-CoV-2 ^14^. In the present work we test the utility of this model in predicting vaccine efficacy against SARS-CoV-2 variants. We find that in vitro neutralisation remains strongly correlated with protection against SARS-CoV-2 variants and the observed protection is consistent with the predictions of the model (Fig 2). A feature of this model that is likely to be useful for future vaccine development is that it provides not just a predicted efficacy, but also a lower confidence bound on the predicted efficacy against each variant for vaccines of a given potency (measured as neutralisation titre to ancestral virus). It should be noted that all reported vaccine efficacies were above this lower bound (Figure 2). However, while the model predictions are consistent with observed vaccine efficacy against symptomatic SARS-CoV2 infection (Figure 2a), we have relatively little data on vaccine efficacy against severe SARS-CoV-2 infection and mortality (Figure 2b). Thus, while vaccine protection against severe infection is an important question explored in our study, it is also the result for which we have the lowest confidence based on existing data. Interestingly, the results also hint that the design of study may play some role in the observed efficacy level. For example, the four randomised control trials are placed relatively symmetrically around the predictive line (one above and three below the mean prediction line)(Figure 2a). However, all 10 observational test-negative case control (TNCC) studies are placed above the line. It is known that test-negative study designs have a number of potential confounders^55,56^, and our analyses suggest future studies should investigate whether TNCC trials have a tendency to over-estimate vaccine efficacy.

The observed correlation between neutralisation and protective efficacy does not prove that neutralisation is the sole mechanism of protection, particularly since many other immune responses (such as cellular responses) are often highly correlated with neutralisation^57^. In addition, although the observed efficacies fall within the 95% predictive interval of the model, it is notable that neutralisation seems better correlated with protection at high neutralisation levels than at lower levels (Figure 2). The uncertainty in the model at low neutralisation is somewhat matched by uncertainty in the efficacy estimates themselves, arising from the generally smaller size of the RCT of vaccines against VOC (see confidence intervals in Supplementary Figure S4). In addition, it is also possible that neutralising responses are a good predictor of efficacy at high neutralisation levels, but other mechanisms such as cellular immunity may contribute a more substantial role at lower neutralisation levels^58,59^. This suggests that future studies investigating alternative immune correlates of protection should be focused on outcomes in populations immunized with more modestly protective vaccines that induce lower neutralising antibody levels.

The combined effects of waning immunity and reduced recognition of the SARS-CoV-2 VOC suggest that vaccine boosters may be needed to maintain high levels of protection from symptomatic SARS-CoV-2 infection. This raises a number of important questions about the potential benefits of boosting and whether variant-specific immunogens will be required. Our analysis suggests maximising neutralising antibody responses to the ancestral virus, through booster vaccination of previously infected individuals (with ancestral immunogens), should be an effective strategy to broadly increase neutralisation titres against SARS-CoV-2 variants. However, the limited studies of boosting in vaccinated subjects raise a number of important questions. Firstly, the optimal timing of boosting is unclear. Most boosting studies in convalescent or previously vaccinated individuals occurred around 6 months after infection or vaccination. One study comparing early versus late boosting of vaccinees would appear to suggest a benefit in delaying to six months^51^. This is consistent with an observed rise in memory B cells within the first months after infection ^53^. Beyond the first months, van Gils and colleagues found a similar increase in binding antibody levels in subjects infected 1 to 15 months before vaccination^60^. It is also presently unclear whether all vaccines will boost immunity to a similar extent, or whether homologous or heterologous boosting might be preferred or made necessary by anti-vector immunity. However, three doses of a potent mRNA vaccine^50^ appear to produce higher levels of neutralisation compared to three doses of lower potency vaccine^51,61^, suggesting that high vaccine potency is important for maximal boosting (Figure 3). The combined effects of waning immunity and reduced recognition of the SARS-CoV-2 VOC raise concerns that boosting of immunity may be needed to maintain high levels of protection from symptomatic SARS-CoV-2 infection. Although the effects of boosting on efficacy are not yet clear, a recent study has suggested that vaccination of previously infected individuals leads to increased protection compared to vaccination of naïve individuals^62^. This raises a number of important questions about the potential benefits of boosting and whether variant-specific immunogens will be required.

Our study has focused upon the effects of boosting with the ancestral SARS-CoV-2 spike immunogen in individuals initially infected with ancestral virus. However, a likely future approach is to develop variant-specific booster vaccines. A recent study compared variant specific (booster) immunisation with a beta (B.1.351) variant SARS-CoV-2 spike construct compared to using the existing vaccine carrying the ancestral strain^50^. Immunisation with a beta spike immunogen led to greater boosting of the beta-specific response (by a further 10% relative to boosting with the ancestral strain)^50^, but at the cost of reduced boosting of responses to the ancestral and gamma strains (which gained only 50-60% of the boost in recognition that was achieved using the ancestral strain). This suggests that immune imprinting or cross reactivity to previous immunogens may need to be considered in strategies aimed at boosting of neutralising responses to new variants ^7^. Additional complexities arise when considering that most current infections are occurring with VOC, rather than the ancestral SARS-CoV-2 virus. Recent studies of neutralising antibody responses after infection with the either the alpha (B.1.1.7)^7^ or beta (B.1.351)^63^ or gamma (P.1)^7^ SARS-CoV-2 variants has shown that infection with these variants induced cross-reactive antibodies against the ancestral strain and other variants. However, in some cases a much greater loss of neutralisation was observed. Keeping an overall high magnitude of the response through boosting may be both an effective and sufficient strategy to maintain neutralisation of variants, even in the absence of variant-specific immunogens. Separate from the effects of boosting on immune protection, the ethical challenges of providing a third vaccine dose to selected populations while others are yet to receive any vaccination are considerable^64^.

Further standardisation and validation of predictive markers of SARS-CoV-2 immunity are urgently needed to allow comparisons of immune responses to current and emerging SARS-CoV-2 variants. In addition, a better understanding of the parameters around the longevity, cross reactivity, and boosting of immunity to SARS-CoV-2 is required to support the development of next-generation and variant-specific vaccines. However, in the absence of such novel vaccine platforms, our analysis suggests that maximising neutralising antibody responses to the ancestral virus using both highly potent initial regimens and through boosting as responses decline may be an effective interim strategy to provide protection from SARS-CoV-2 variant infection.

## Supporting information

Supplemental Analysis and Methods

## Data Availability

All data and code will be made freely available on GitHub upon publication.

## Ethics statement

This work was approved under the UNSW Sydney Human Research Ethics Committee (approval HC200242).

## Authorship Statement

All authors contributed to the data collection, design of the study, writing of the manuscript and revision of the manuscript. DSK, DC, AR, TES and MPD contributed to the modelling and statistical analysis of the data.

## Data Availability Statement

All data and code will be made freely available on GitHub upon publication.

## Acknowledgements

This work would not be possible without the many scientists who generously provided the published data analysed in this study, either through making the data directly available through the original publication or through providing it upon request. The authors thank these scientists for their contribution and the individual sources of data are indicated in the references and supplementary tables.

## Supplementary figures

**Figure S1:**
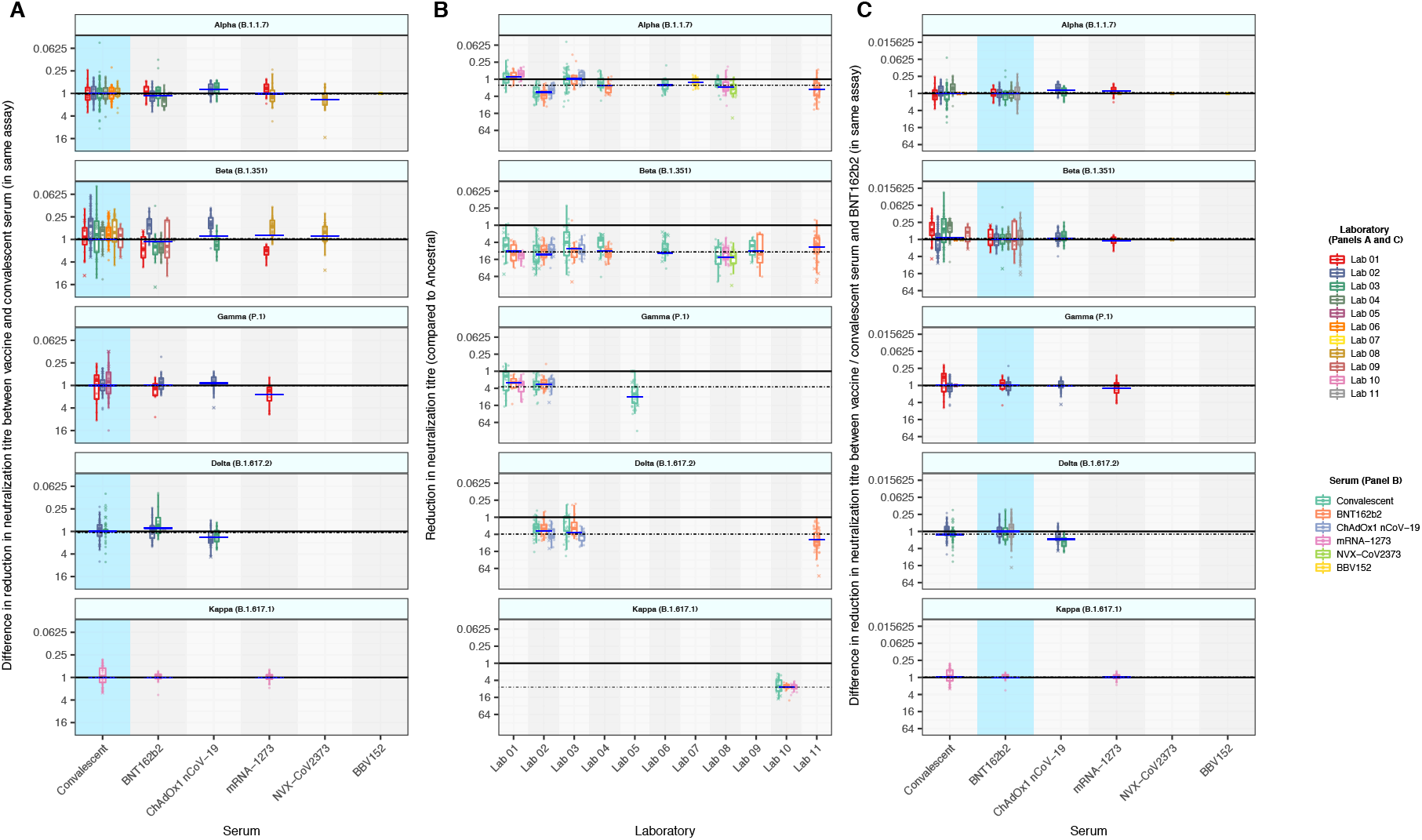
In vitro neutralisation of SARS-CoV-2 variants. (A) Change in neutralisation titre of all vaccinees normalised against the change in neutralisation titre seen in convalescent individuals in the same study. For convalescent subjects, the mean for each study is one (since titres are normalised to convalescent). For different vaccination groups, the difference between the drop in titre in convalescent individuals in the same study is shown. Horizontal bars indicate the weighted mean for that vaccine / variant combination. Vaccination groups show changes in neutralisation titre that closely match that of convalescent subjects in the same study. (B) For each laboratory the mean change in neutralisation titre observed in convalescent subjects and different vaccine groups is shown. Although estimates of change in neutralisation vary between laboratories, within a given laboratory the change in neutralisation titre is congruent between convalescent and different vaccine groups. (C) Normalisation against BNT162b2 vaccinee sera; Panel A normalises vaccine responses against convalescent sera (which are not consistently defined across different studies). To check that our conclusions are robust to the reference serum used, we also analysed the subset of studies in which sera from individuals vaccinated with BNT162b2 was available and normalised against the change in neutralisation titre seen in BNT162b2-vaccinated subjects. As can be seen, the dominant effect of laboratory is still evident when normalised against BNT162b2 sera.

**Figure S2:**
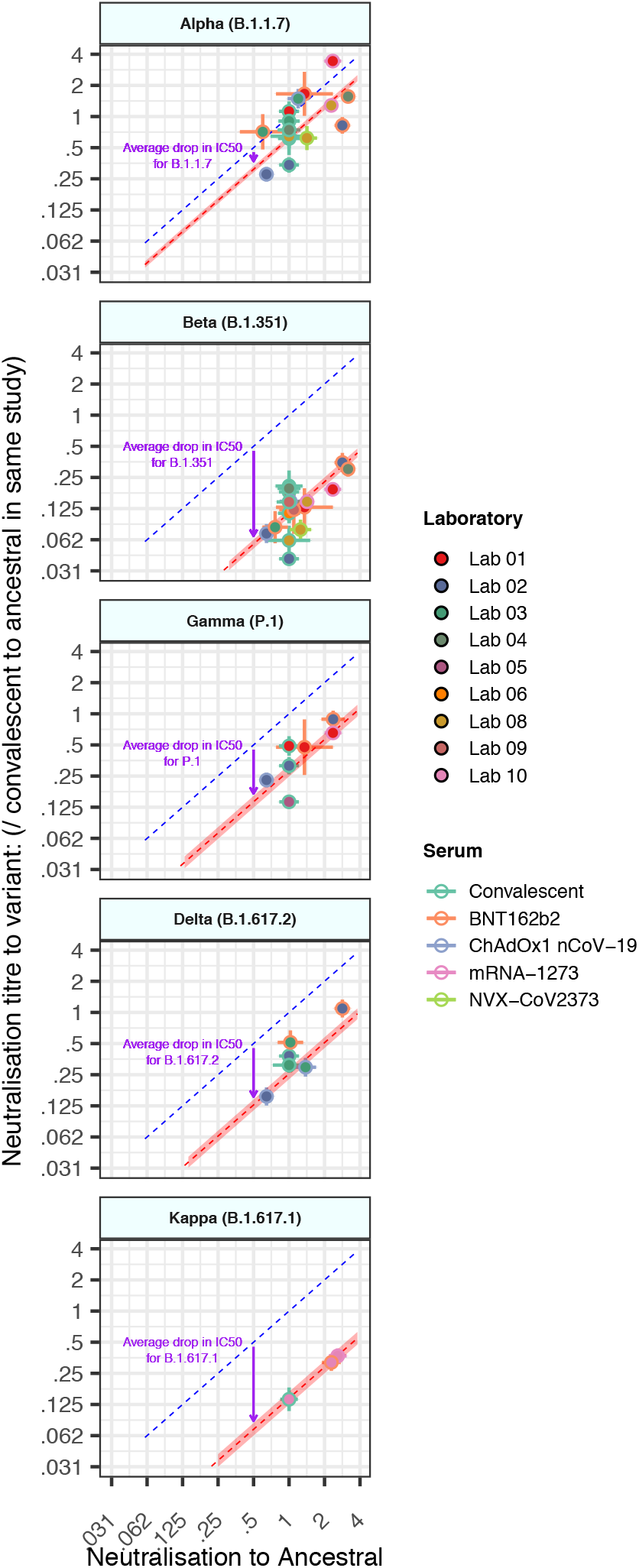
Neutralisation of ancestral virus predicts neutralisation of variants. The mean neutralisation titre against the ancestral virus (x-axis) and the mean neutralisation titre against the VOC (y-axis) is shown for individual studies. The predicted line for a 1:1 relationship is indicated (dashed blue line). The observed mean drop in neutralisation titre across all vaccines and convalescent subjects is indicated by an arrow, and the predicted levels of variant neutralisation are indicated by a dashed red line (shading indicates 95% CI) are shown. The results for individual studies are variable because of differences between assays and Figure 2 reports the mean across all studies for each vaccine / variant combination.

**Figure S3:**
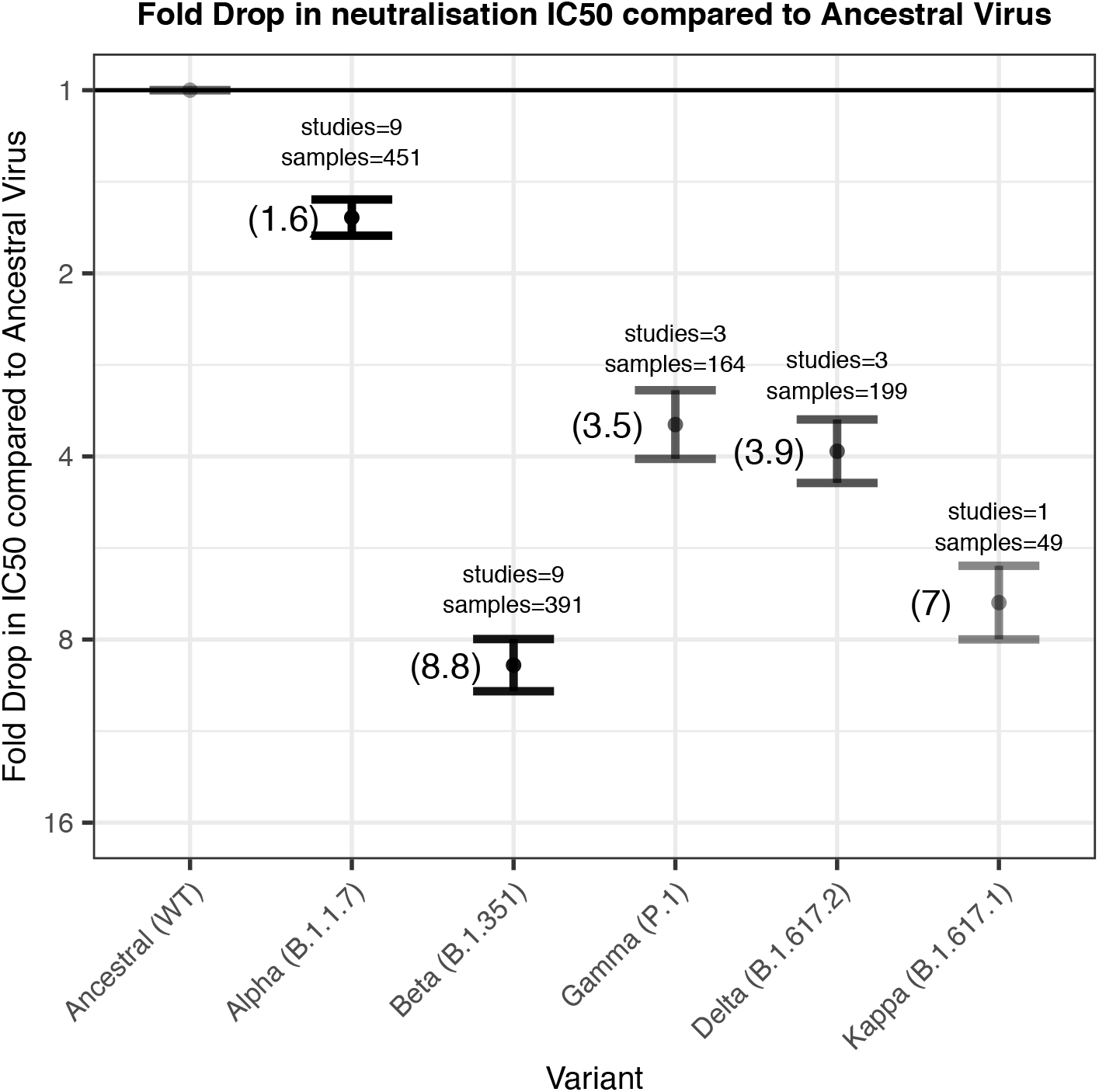
Mean drop in neutralisation titre against SARS-CoV-2 variants. The mean fold-drop in neutralisation titre reported for different SARS-CoV-2 variants is shown (with 95% CI). The number of subjects and studies contributing to this is also indicated.

**Figure S4:**
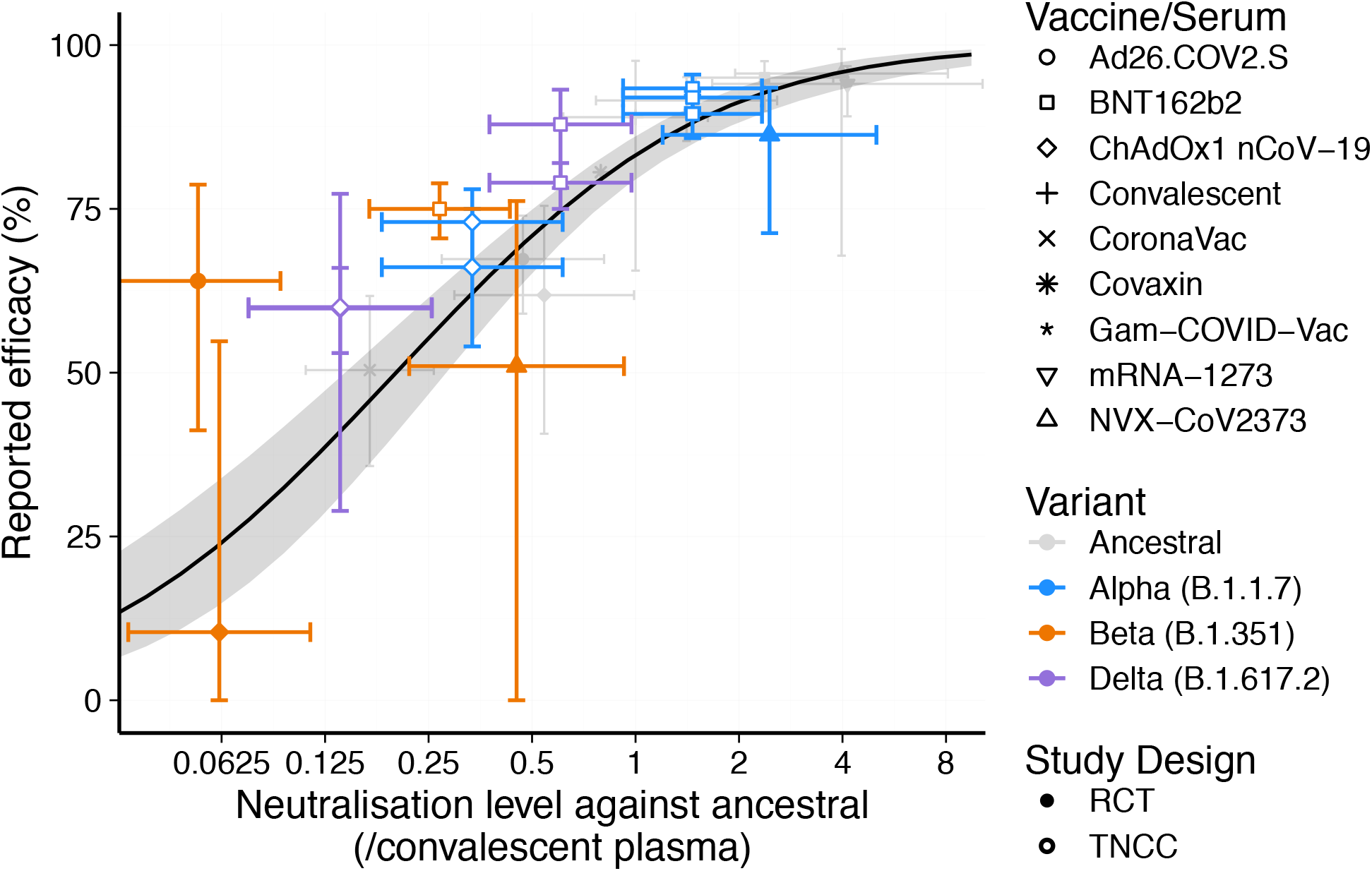
Correlation between in vitro neutralisation and observed protection. The relationship between neutralisation and protection derived from data on ancestral virus is shown (mean as solid line, shading is 95% CI). The predicted in vitro neutralisation titres against variants (based on the titres reported against ancestral virus in phase I/II studies, and adjusted for the mean drop in neutralisation titre to variants reported in supplementary Figure S3) are shown for each vaccine, along with the observed efficacy against VOC (see supplementary table 2). Note that whereas in Figure 2 the model curve is adjusted by the mean drop in neutralisation to VOC, here the mean neutralisation titres for each vaccine / variant combination are adjusted for this drop.

