## Supplemental Analysis and Methods for "SARS-CoV-2 variants: levels of neutralisation required for protective immunity"

**Supplementary analysis**

*Analysis of data on neutralisation against variants*

In this study we aimed to determine the factors that influenced the loss of recognition against SARS-CoV-2 variants of concern (VOC), and more specifically, whether different vaccines varied in their recognition of VOC. The loss of recognition of vaccines to VOC is typically measured as the 'drop in neutralisation titre' against the variants (ie: the change in the concentration of serum needed for 50% neutralisation in vitro). Therefore, we obtained data from 16 published studies which directly compared neutralisation titre against ancestral (Wuhan-like / D614G strains) and the VOC. This included data provided in the original publications or sent by authors (see Supplementary table 1). We focused primarily on assays using live SARS-Cov-2 virus (to reduce the potential variability that might arise from different pseudoviral constructs[1]), with the exception of the NVX-CoV2373 vaccine, for which only data from neutralisation assays using a spike-expressing pseudovirus was available.

When combining data from multiple studies, an important caveat is that different laboratories used distinct in vitro assays to measure neutralisation of SARS-CoV-2 (Supplementary table 1) [1]. These assays differ considerably in the mean neutralisation titres and fold-change to variants reported, even when considering notionally similar groups of subjects, such as convalescent serum against the ancestral virus (Figure 1a). To test the extent to which a given vaccine platform affects antibody cross reactivity, we calculated the mean drop in neutralisation titre across different vaccines and variants (comparing to ancestral virus), while accounting for censoring at the assay specific limit of detection (censoring to estimate means and in regression analysis is described in supplementary methods). We found a large variation in drops in titres in vaccine serum between variants, vaccines and laboratories. For example, comparing 7 studies reporting the change in neutralisation titre of vaccine sera against the beta (B.1.351) variant (compared to ancestral virus) following vaccination, the estimated decrease in neutralisation titre ranges from 6-fold[2] to 16-fold[3], depending on the study and the vaccine considered. Superficially, these differences might suggest the vaccines elicit antibody responses with different levels of cross-reactivity to the VOC. However, many studies also included a direct comparison of convalescent sera or included sera from different vaccines. When the change in neutralisation titre in vaccinees is compared with the change seen in convalescent subjects in the same study, this is usually very similar (Figure S1a, b).

The same is true when using a vaccine comparator group instead of convalescent subjects (Figure S1c).

The variability in results between assays means that we observe several estimates for the extent of antibody cross-reactivity against a given variant (Figure 1a). It is not clear if one assay is more accurate than another in capturing the biologically relevant change in neutralisation. Therefore, we estimated the mean drop in neutralisation titre for a given variant across the available studies by aggregating all individual data (convalescent and vaccine) from each study, and using censoring to account for the differing limit of detections of each assay (supplementary methods). We find the mean drop in neutralisation titre is 1.6-fold for the alpha/B.1.1.7 variant (95% CI = 1.5 -1.7, average of n= 9 studies), 8.8-fold (95% CI = 8 -9.7, n= 9 studies) for the beta/B.1.351 variant, 3.5-fold (95% CI = 3.1 - 4, n= 3 studies) for the gamma/P.1 variant and 3.9-fold (95% CI = 3.5 - 4.4, n= 3 studies) for the delta/B.1.617.2 variant (Figure S3).

We then performed censored regression to assess the impact of vaccine type on cross-reactivity. We accounted for both variant and laboratory specific effects. The variant specific effect was incorporated by including the mean drop in neutralisation titre seen between ancestral virus and each variant virus across all studies as described above ( $\Delta_V$). A laboratory specific effect was incorporated by including a factor for each laboratory ($L$). We also allowed for an additional categorical variable for serum type to be included ($S$). $S$ was a factor that determined whether the serum came from a convalescent or vaccinated individual, and if it was the latter, which vaccine was used. Thus the model was:

$$Variant\ Neut \sim \alpha_0 + \alpha_1 Ancestral\ Neut + \Delta_V + \alpha_2 L + \alpha_3 S \qquad \text{(Eq S1)}$$

We found that in this model laboratory was a highly significant factor (p<.0001), but after including laboratory, serum type (S) was not a significant factor in the model (p=0.256) (detailed in supplementary methods). Additionally, the best model (as judged by the model with the lowest AIC was one that included $L$ but not $S$. This does not mean that vaccine sera all neutralise VOC equally. That is, vaccinee sera vary considerably in their ability to neutralise ancestral virus. However, they all tend to *lose recognition* of a given VOC to a similar extent (ie: comparing the drop in titre between ancestral and VOC). This demonstrates that the neutralisation titre of ancestral virus is a very good predictor of neutralisation of a variant (Figure 1b, S1a, S2), and that once the initial neutralisation level and variation between labs is considered the vaccine platform itself is not a factor in the model.

We additionally performed censored regression to determine the best predictors of the *drop* in neutralization against a variant (compared to ancestral virus) for each vaccine used. For this we modelled

$$Fold\ Drop_v \sim \alpha_0 \Delta_V + \alpha_1 L + \alpha_2 \mu_{conv_L} + \alpha_3 S$$

(Eq S2)

Where $\mu_{conv_L}$ was defined as the mean fold-drop in convalescent sera against each variant for each laboratory, $L$ was once again a categorical factor for the laboratory and $S$ was a categorical factor for the serum used (however here only vaccinated individuals were considered in the model, as convalescent individuals were used to normalise the fold-drop by laboratory). For this model we allowed only one of $\alpha_1$ or $\alpha_2$ to be non-zero (so that there was only one laboratory specific effect). We found that once again, after accounting for laboratory specific effects, the vaccine used was not a significant factor, regardless of whether $\alpha_1$ or $\alpha_2$ (or neither) were included in the model (p>.07 in all cases, likelihood ratio test). Additionally, we found that the best model (i.e. the model with the lowest AIC) was one that included $\mu_{conv_L}$but did not include $L$ or $S$. This means that a combination of (i) the mean drop in neutralisation titre seen between ancestral virus and each variant virus across all studies and (ii) the mean drop in neutralisation titre seen in convalescent sera in the current study are the best predictors of the fold drop in neutralisation titre for the serum under consideration.

Together, these results suggests that any one study of neutralisation of a variant does not provide a reliable estimate of the cross-reactivity of serum to variants, but the best estimate of neutralisation against current VOC is obtained from the neutralisation observed against the ancestral virus, combined with the average fold drop in neutralisation to the particular variant observed across multiple studies.

*Estimating neutralisation after boosting*

To estimate the neutralisation level achieved in individuals after boosting, we used data from studies of boosting of previously infected individuals or previously vaccinated individuals. Studies were included if they contained a comparison with naïve vaccinated individuals within the same study or if a similar comparison of vaccination in naïve individuals could be made with a separate study by the same laboratory using the same assay (detailed in Supplementary table 4). This allowed us to determine the fold increase in neutralisation level in previously infected/vaccinated individuals compared with naive individuals who received the standard two-dose vaccine regimen. All boosted individuals received either BNT162b2, mRNA-1273 or CoronaVac. To calculate the neutralisation level in individuals after boosting (Figure 3), the neutralisation level reported in naïve individuals in the phase I/II trials for each vaccine (as reported in the Supplementary table S3 from reference[4]) were multiplied by the fold increase reported in the boosting studies in Supplementary table 3. For some studies – naïve or previously infected individuals were reported to have received either BNT162b2 or mRNA-1273 - and data on which individuals received which vaccines was not paired with the neutralisations titres [5-7]. In these cases the geometric mean of the neutralisation reported in the phase I/II trials for BNT162b2 and mRNA-1273 was calculated (geometric mean: 3.13 fold of convalescent plasma) and the fold-increase between vaccination in previously infected individuals and naïve individuals was multiplied by this geometric mean level for naïve individuals receiving one of the two vaccines. This provided a range of estimates of the neutralisation level (as a fold of convalescent plasma) after boosting of previously infected individuals of between 6.1-28.7, with geometric mean of 12.0 (red shaded region and dashed line, Figure 3).

*Estimating decay in vaccine efficacy*

In this study we aimed to estimate the efficacy against variants over the first year (with and without boosting). Modelling the decay in efficacy was performed by determining (from the inverse of the model in Equation S8 below), the neutralisation titres expected to give an initial target efficacy (ie: 95%, 90%, 80% or 70%) against infection with the ancestral virus. Neutralisation was assumed to decay over the first 360 days with a half-life of 108 days (as estimated in [4], using data from [8]). The efficacy at each time point was then determined from the neutralisation level after decay until that time point (using Equation S8). Neutralisation to variants was assumed to be reduced by the same fold change (Figure S3), as titres decayed. Boosting was modelled as an increase in the neutralisation level to the mean level determined from studies in previously infected or vaccinated individuals (red dashed line in Figure 3), assuming decay rate in neutralisation was the same after boosting as before, and assuming that loss of neutralisation to variants was the same fold-change as prior to boosting. The lower bound on efficacy estimates from the model was determined using the bootstrapping approach described in the supplementary methods.

**Supplementary Methods**

In this section we describe the methods used to estimate the average fold-drop in neutralisation against each variant and how this was used to predict the efficacy of vaccines against each variant. A major focus of these methods is in accounting for censoring of neutralisation measurements when they fell below the limit of detection, and standard censoring models cannot be used because different assays had different limits of detection. Another major focus of these methods is to explain how bootstrapping was used to determine the error in predictions of vaccine efficacy.

To aid reading of these methods it is worth noting that throughout these methods we will use subscripts to refer to the serum type under consideration and superscripts to refer to the virus type under consideration. A superscript of either *a* or *0* refers to the special case of ancestral virus.

The following letters are paired with these subscripts and superscripts: (i) Neutralisation titre for serum *i* against variant *v* is depicted by $N_i^v$; (ii) Fold change in neutralisation titre from variant $v_j$ to variant *v* for serum *i* is depicted by $F_i^{v,v_j}$. Where variant $v_j$ is ancestral virus (i.e. $F_i^{v,a}$) this is shortened to $F_i^v$; (iii) The limit of detection for serum *i* against variant *v* is depicted by $L_i^v$; (iv) Left and right censoring variables for serum *i* against variant *v* are denoted by $c_{L_i}^v$ and $c_{R_i}^v$, respectively.

*Estimating the mean fold-change in neutralisation against each variant with censoring at the limit of detection*

When estimating the mean fold-change in sera neutralisation of ancestral virus versus a SARS-CoV-2 variant ($F_i^v$) it was important to adjust for the censoring of data when neuralisation against the variant or ancestral virus fell below the limit of detection. When the neutralisation titres for a serum sample against the ancestral virus ($N_i^a$) and a variant ($N_i^v$) were both above the limit of detection the fold-change in neutralisation was calculated as $F_i^v = \frac{N_i^v}{N_i^a}$. When the neutralisation titre declined from a value above the limit of detection against the ancestral virus, to below the limit of detection against the variant ($L_i^v$), the fold-change in neutralisation was $F_i^v \leq \frac{L_i^v}{N_i^a}$ (this occurred in 98 samples). In this case we set the left censoring variable, $c_{L_i}^v$ to be 1. In the uncommon case (2 samples) where the neutralisation against the ancestral virus was below the $(L_i^a)$, but the neutralisation against the variant was above the limit of detection the fold change was $F_i^v \geq \frac{N_i^v}{L_i^a}$, and this possibility accounts for the times when there is a detected increase in neutralisation titre against variant compared with ancestral. In this case we set the right censoring variable, $c_{R_i}^v$ to be 1. In all other cases the left and right censoring variables were set to 0. When the neutralisation titre against the ancestral virus and variant were below the limit of detection these data were excluded as they provided no information on neutralisation change (29 samples). To estimate the mean fold-change in neutralisation against a particular variant $(v)$ we assumed a normal distribution for the log-transformed fold changes observed in sera samples against that variant, (i.e. $\log_{10} F_i^v$), and fitted this using maximum likelihood estimation. The likelihood function for fitting this normal distribution (including censoring) was,

$$\mathcal{L}_v(\boldsymbol{F^v}, \boldsymbol{N^v}, \boldsymbol{N^a}, \boldsymbol{L^v}, \boldsymbol{L^a}\boldsymbol{C_L^v}, \boldsymbol{C_R^v} \mid \mu^v, \sigma^v) =$$

$$\prod_{i=1}^{S^v} g(\log_{10}(F_i^v), \mu^v, \sigma^v)^{1-c_{L_i}^v - c_{R_i}^v} \, G\left(\log_{10}\left(\frac{L_i^v}{N_i^a}\right), \mu^v, \sigma^v\right)^{c_{L_i}^v}$$

$$\left(1 - G\left(\log_{10}\left(\frac{N_i^v}{L_i^a}\right), \mu^v, \sigma^v\right)\right)^{c_{R_i}^v}$$

(Eq S3)

where the i[th] elements of the vectors $\boldsymbol{F^v}, \boldsymbol{N^v}, \boldsymbol{L^v}$, $\boldsymbol{C_L^v}$ and $\boldsymbol{C_R^v}$ vectors are $F_i^v$, $N_i^a, N_i^a$, $L_i^v$, $L_i^a, c_i^L$ and $c_i^R$, respectively. The function $g(x, \mu, \sigma)$ is the probability density at $x$ of a normal distribution with mean $\mu$ and standard deviation $\sigma$. The function $G(x, \mu, \sigma)$ is the cumulative density at $x$ of a normal distribution with mean $\mu$ and standard deviation $\sigma$. The mean $(\mu^v)$ and standard deviation $(\sigma^v)$ of the normal distribution that minimise the negative log of this likelihood function were found using the *nlm* function in the R statistical package (version 4.0.2).

*Creating a censored regression model to predict neutralisation against variant virus*

In order to predict neutralisation titre against variant virus using the multiple regression model in Equation S1, we set up a custom censored regression model. The purpose of this custom model is to allow for both neutralisation titre against the variant virus and neutralisation titre against the ancestral virus to potentially be below the limit of detection for a study. Existing censored regression packages (e.g. *CensReg*[9]) were not used as the limit of detection was different for different laboratories – though *CensReg* produced similar results in a parallel analysis.

We set up a likelihood function to estimate the neutralisation against each variant using the neutralisation against ancestral virus. We considered $n_v$ different variants, and, using Equation S3 we estimated the mean fold change in neutralisation titre against each variant (compared to ancestral virus) across all serum samples available, and denoted this, $\bar{F}^v$. Here we denote ancestral virus by $v=0$. The vector of these fold changes across all variants is denoted $\overline{\boldsymbol{F}}$. We also stored the neutralisation titres and limits of detection for each variant / serum (including convalescent serum) combination in matrices $\boldsymbol{N}$ and $\boldsymbol{L}$ respectively, with $\boldsymbol{N} = (N_i^v)$ and $\boldsymbol{L} = (L_i^v)$. For each serum sample we recorded the study ($A_i$) and serum type ($T_i$) and stored these in vectors $\boldsymbol{A}$ and $\boldsymbol{T}$ respectively. Finally, the lower and upper bounds for censoring (due to either variant neutralisation being below the limit of detection or ancestral virus neutralisation titre being below the limit of detection, as described above) were stored in matrices $\boldsymbol{C_L} = \left(C_{L_i}^v\right)$ and $\boldsymbol{C_R} = \left(C_{R_i}^v\right)$, respectively. We define a vector of parameter estimates $\boldsymbol{\alpha}$ as $\boldsymbol{\alpha} = (\alpha_0, \alpha_1, \alpha_2, \alpha_3, \alpha_4)$; corresponding to the coeffcients in the regression model.

The likelihood function (including censoring) that we used was

$\mathcal{L}_v(\boldsymbol{N}, \overline{\boldsymbol{F}}, \boldsymbol{L}, \boldsymbol{A}, \boldsymbol{T}, \boldsymbol{C_L}\boldsymbol{C_R}|\, \boldsymbol{\alpha}, \sigma) =$

$$\prod_{v=1}^{n_v}\prod_{i=1}^{S^v} g(\log_{10}(N_i^v), h_i^v, \sigma^v)^{1-C_{L_i}^v-C_{R_i}^v}\, G(\log_{10}(L_i^v\,), h_i^v, \sigma^v)^{C_{L_i}^v}$$

$$\left((1 - G(\log_{10}(L_i^0), h_i^v, \sigma^v)\right)^{C_{R_i}^v}.$$

(Eq S4)

Where $h_i^v$ is defined as:

$$h_i^v = \alpha_0 + \alpha_1 N_i^0 + \bar{F}^v + \alpha_2 A_i + \alpha_3 T_i.$$

(Eq S5)

We minimised the negative log of the likelihood function in Equation S4 for models where neither, one or both of $\alpha_3$ and $\alpha_4$ were set to zero using the *optim* function in the *stats* package of R (version 4.0.2). We then compared models using the likelihood ratio test.

*Creating a censored regression model to predict fold drop in neutralisation titre for variant virus*

In a similar way to that described above, we set up a censored regression model to predict the fold change in neutralisation titre for each variant. In this case, and using the same notation outlined above, the likelihood function (including censoring) that we used was

$$\mathcal{L}_v(\boldsymbol{F},\overline{\boldsymbol{F}},\boldsymbol{L},\boldsymbol{A},\boldsymbol{T},\boldsymbol{C_L}\boldsymbol{C_R}|\,\boldsymbol{\alpha},\sigma) =$$

$$\prod_{v=1}^{n_v}\prod_{i=1}^{S^v} g(\log_{10}(F_i^v),k_i^v,\sigma^v)^{1-C_{L_i}^v-C_{R_i}^v}\, G\left(\log_{10}\left(\frac{L_i^v}{N_i^0}\right),k_i^v,\sigma^v\right)^{C_{L_i}^v}$$

$$\left((1-G\left(\log_{10}\left(\frac{N_i^v}{L_i^0}\right),k_i^v,\sigma^v\right)\right)\right)^{C_{R_i}^v}.$$

(Eq S6)

Where $k_i^v$ is defined as:

$$k_i^v = \alpha_0 + \bar{F}^v + \alpha_1\overline{FC_{A_i}^v} + \alpha_2 A_i + \alpha_3 T_i.$$

(Eq S7)

and $\overline{FC_{A_i}^v}$ represents the average fold drop in convalescent sera for variant *v* in the lab from which serum *i* comes. As in Equation S2, at most one of $\alpha_1$ or $\alpha_2$ were allowed to be non-zero.

*Predicting the efficacy for variants based on previously developed model*

Previously, we developed and fitted a model of vaccine efficacy to data on the immunogenicity and protective efficacy (against symptomatic and severe COVID-19) of 7 vaccines from phase I/II and phase III trials, respectively [4]. Here we use this model, as originally published and parameterised, to predict the efficacy of vaccines against each variant, using the fold-change in neutralisation titre estimated against each variant in this study (Figure S3). The model estimates protective efficacy of a vaccine as,

$$P(n_{50},k,\mu_s^v,\sigma_{all}) = \int_{-\infty}^{\infty} E(n \mid n_{50},k)\, f(n \mid \mu_s^v,\sigma_{all})\ dn$$

(Eq S8)

where $\mu_s^v$ is the ($\log_{10}$) mean neutralisation titre of a vaccine ($s$) against variant ($v$) (normalised to the mean of convalescent sera against ancestral virus), $\sigma_{all}$ is the standard deviation in the neutralisation titres across individuals, $f$ is the probability density of a normal distribution with mean $\mu_v$ and standard deviation $\sigma_{all}$, and $E$ is a logistic function of the form,

$$E(n \mid n_{50}, k) = \frac{1}{1 + e^{-k(n-n_{50})}}. \qquad \text{(Eq S9)}$$

The parameter $n_{50}$ is the ($\log_{10}$) neutralisation level that provides an individual with 50% protective efficacy of COVID-19, and $k$ is the parameter determining the steepness of the logistic relationship. A number of these parameters were estimated previously for symptomatic and severe COVID-19 (Supplementary table 4) [4].

To investigate the ability of the previously published model to predict vaccine efficacy against variants, we compared our models prediction of vaccine efficacy with the observed efficacy for each variant and vaccine combination we identified in the literature (Figure 2). To estimate the mean neutralisation level of each vaccine against each variant (where efficacy data was available), we calculated

$$\mu_s^v = \mu_s + \bar{F}^v$$

where $\bar{F}^v$ is the ($\log_{10}$) mean fold-change in neutralisation titres for each variant (calculated above as above) and $\mu_s$ is the neutralisation level reported for each vaccine (ratio of neutralisation titre in vaccinated individuals compared with convalescent individuals) that was reported in Phase I/II trials against ancestral virus [4] (Figure 2).

*Determining the confidence and lower bound of predicted efficacy using parametric bootstrapping*

During vaccine development it is useful to know the uncertainty in efficacy predictions as measured by the confidence interval for the efficacy estimate. In particular, the lower confidence bound for efficacy for a given neutralisation level is useful as an estimate of the minimum expected level of achieved efficacy (Figure 2). Confidence intervals (and lower bounds) of predicted efficacies (shaded regions) in Figures 2, 4 and S4 were generated using parametric bootstrapping on the parameters with uncertainty in their estimation (Supplementary table 4) as follows. For any neutralisation ratio (i.e. position on the x-axis in Figure 2 and S4), Equation S8 was first used to estimate the mean corresponding protective efficacy against a particular variant. Then the distribution of likely efficacies was estimated by repeating the efficacy calculation with Equation S8, using parameter values chosen randomly from distributions according to their standard error or covariance matrix (normal and bivariant normal distributions respectively, Supplementary table 4). The sources of uncertainty include the model parameter uncertainty estimated in the previous study [4], as well as the uncertainty in estimates of the neutralisation level and the fold drop in neutralisation to each variant (Figure S3). It should be noted that when estimating the neutralisations level for each vaccine there is between laboratory uncertainty (i.e. due to differences in laboratory and assays used), and within laboratory uncertainty. The between laboratory variability in estimates of neutralisation level (standard error in estimates 0.18) was determined in this study and found to be less than the maximum within laboratory uncertainty (standard error in estimates, 0.20). Therefore, we used the largest within study uncertainty as the measure of uncertainty in the efficacy estimates (Supplementary table 4). The distribution in efficacy was generated from 10,000 bootstraps for each neutralisation level and the 95% confidence limits estimated using the percentile method (2.5 and 97.5 percentile for a regular 95% confidence interval, Figure 2, 4 and S4).
