## Supplemental Tables 1-4 for "SARS-CoV-2 variants: levels of neutralisation required for protective immunity"

### Supplementary Table 1:

| Reference | Serum type | Dosage | Measured on day | Assay | Variants tested | Data derived from: | Figure Lab Number |
|---|---|---|---|---|---|---|---|
| 1 | Convalescent | n/a | > 1 month post recovery (severe and non-severe) | Live virus end point microneutralisation assay, ID50 | Ancestral (WA1)<br>Alpha/B.1.1.7<br>Beta/B.1.351 | Fig 3b and Fig 4b, raw data kindly provided | Laboratory 1 |
| | mRNA-1273 | 100 µg | 15 days post second dose | | | | |
| | BNT162b2 | ns | > 7 days post second dose | | | | |
| 2 | Convalescent | n/a | > 1 month post recovery (severe and non-severe) | Live virus end point microneutralisation assay, ID50 | Ancestral (WA1)<br>Gamma/P.1 | Fig S1C and D, raw data kindly provided | Laboratory 1 |
| | mRNA-1273 | 100 µg | 15 days post second dose | | | | |
| | BNT162b2 | | >7 days post second dose | | | | |
| 3 | Convalescent | n/a | 4-9 weeks post infection | Live virus focus reduction neutralisation test, FRNT50 | Ancestral (Victoria)<br>Beta/B.1.351 | Fig 2c/Table S1a | Laboratory 2 |
| | BNT162b2 | 30 µg | 7-17 days post second dose | | | Fig 3c/Table S2 | |
| | CHAdOx1/AZD1222 | Standard or half dose | 14 or 28 days post second dose | | | Fig 3d/Table S2 | |
| 4 | Convalescent | n/a | 4-9 weeks post infection | Live virus focus reduction neutralisation test, IC50 | Ancestral (Victoria)<br>Gamma/P.1 | Table S4A | Laboratory 2 |
| | BNT162b2 | 30 µg | 4-17 days post second dose | | | Table S5 | |
| | CHAdOx1/AZD1222 | Standard or half dose | 14 or 28 days post second dose | | | | |
| 5 | Convalescent | n/a | 4-9 weeks post infection | Live virus focus reduction neutralisation test FRNT50 | Ancestral (Victoria)<br>Alpha/B.1.1.7 | Fig5a, raw data kindly provided | Laboratory 2 |
| | BNT162b2 | 30 µg | 7-17 days post second dose | | | Fig5b, raw data kindly provided | |
| | CHAdOx1/AZD1222 | Standard dose (5 x $10^{10}$) or half dose | 14 or 28 days post second dose | | | Fig5c, raw data kindly provided | |
| 6 | Convalescent | n/a | 4-9 weeks post infection | Live virus focus reduction neutralisation test, FRNT50 | Delta/B.1.617.2 | Table S4 | Laboratory 2 |
| | BNT162b2 | 30 µg | 7-17 days post second dose | | | | |
| | CHAdOx1/AZD1222 | Standard or half dose | 14 or 28 days post second dose | | | | |
| 7 | Convalescent | n/a | 168-197 days post symptom onset.<br>Mild/moderate, severe and critical disease | Live virus, GFP-split reporter system, IC50 | D614G (hCoV-19/France/GE1973/2020)<br>Alpha/B.1.1.7<br>Beta/B.1.351 | Fig 2b, raw data kindly provided | Laboratory 3 |
| 8 | Convalescent | n/a | 6 months post symptom onset | Live virus, GFP-split reporter system, IC50 | D614G (hCoV-19/France/GE1973/2020)<br>Alpha/B.1.1.7<br>Beta/B.1.351<br>Delta/B.1.617.2 | Fig 4a, raw data kindly supplied | Laboratory 3 |
| | BNT162b2 | ns | Ave 36 days (32-46) | | | | |
| 9 | Convalescent | n/a | ~ 1 month post mild infection | Live virus focus reduction | Ancestral WA1with D614G (recombinant)<br>Alpha/B.1.1.7- isolate | Fig 2a,e, raw data kindly provided | Laboratory 4 |

| | | | | | | | |
|---|---|---|---|---|---|---|---|
| | BNT162b2 | ns | 7 days post second dose | neutralisation test, EC50 | Beta/B.1.351 spike (recombinant) | Fig 4a,d, raw data kindly provided | |
| 10 | Convalescent | n/a | 10-102 days post infection | Live virus Plaque reduction neutralisation test PRNT50 | Ancestral<br>Gamma/P.1 | Supplementary table 4 | Laboratory 5 |
| 11 | Convalescent | n/a | 6-43 days post symptom onset/first PCR test positive | Live virus focus reduction neutralisation test, IC50 | Ancestral (hCoV-19/England/02/2020)<br>Alpha/B.1.1.7<br>Beta/B.1.351 | 2b, raw data kindly provided | Laboratory 6 |
| 12 | BBV152 | ns | ns | Live virus Plaque reduction neutralisation test PRNT50 | Ancestral<br>Alpha/B.1.1.7 | Fig 1b | Laboratory 7 |
| 13 | Convalescent | n/a | 1 to 8 weeks after resolution of infection or 2 to 10 weeks after the most recent positive SARS-CoV-2 test. | Lentivirus based pseudovirus assay, ID50 | Ancestral with D614G<br>Alpha/B.1.1.7 | Fig 2a/Table S1 | Laboratory 8 |
| | mRNA-1273 | 100 µg | 28 days post second dose | | | | |
| | NVX-CoV2373 | 5 µg protein + Matrix M | 14 days post second dose | | | | |
| 14 | Convalescent | n/a | 1 to 8 weeks after resolution of infection or 2 to 10 weeks after the most recent positive SARS-CoV-2 test. | Lentivirus based pseudovirus assay, ID50 | Ancestral with D614G<br>Beta/B.1.351 | Fig 1a/Table S2 | Laboratory 8 |
| | mRNA-1273 | 100 µg | 28 days post second dose | | | | |
| | NVX-CoV2373 | 5 µg protein + Matrix M | 14 days post second dose | | | | |
| 15 | Convalescent | n/a | 2-85 days post + PCR test | Live virus focus reduction neutralisation test, ID50 | B.1.126 ('WT')<br>Beta/B.1.351 | Fig 4a | Laboratory 9 |
| | BNT162b2 | ns | 7-16 days post second dose | | | | |
| 16 | Convalescent | n/a | 31-91 days post symptom onset | Live virus focus reduction neutralisation test, FRNT50 | Ancestral (WA1)<br>Kappa/B.1.617.1 | Fig 1a/Supplementary table 2 | Laboratory 10 |
| | BNT162b2 | ns | 7-27 days post second dose | | | Fig 1c/ Supplementary table 3 | |
| | mRNA-1273 | ns | 35-51 days post second dose | | | Fig 1b/ Supplementary table 4 | |
| 17 | BNT162b2 | ns | Median: 28 days post second dose | Live virus focus reduction neutralisation test, IC50 | Ancestral (hCoV19/England/02/2020)<br>D614G<br>Alpha/B.1.1.7<br>Beta/B.1.351<br>Delta/B.1.617.2 | https://github.com/davidlvb/Crick-UCLH-Legacy-VOCs-2021-05 | Laboratory 11 |

ns. Not specified

### Supplementary Table 2:

| Study | Vaccine | Trial Started (Day) | Variants compared | | | Measure of effectiveness | Efficacy against severe disease | | | Trial design | Data derived from |
|---|---|---|---|---|---|---|---|---|---|---|---|
| | | | Alpha/B.1.1.7 | Beta/B.1.351 | Delta/B.1.617.2 | | Alpha/B.1.1.7 | Beta/B.1.351 | Delta/B.1.617.2 | | |
| 18 | BNT162b2 | 14 days post 2nd dose | 89.5%* | 75%# | N/A | Any documented infection | 100% (95% CI: 81.7-100) | 100% (95% CI: 73.7-100) | N/A | Test negative case control | Table 1 |
| 19 | BNT162b2 | 14 days post 2nd dose | 93.4% | | 87.9% | Symptomatic disease | ND | | | Test negative case control | Table 2 |
| | ChAdOx1 nCoV-19 | 14 days post 2nd dose | 66.1% | | 59.8% | Symptomatic disease | ND | | | | |
| 20 | ChAdOx1 nCoV-19 | 14 days post 2nd dose | | 10.4% | | Symptomatic disease | Not able to report. | | | Randomized controlled trial | Table 2 |
| 21 | NVX-CoV2373 | 7 days post 2nd dose | | 51%^ | | Symptomatic disease | Not able to report | | | Randomized controlled trial | In text |
| 22 | NVX-CoV2373 | 7 days post 2nd dose | 86.3% | | | PCR confirmed symptomatic COVID, mild, moderate or severe | Not specifically reported (only 5 cases of severe disease, all in placebo group) | | | Randomized controlled trial | Fig 4 |
| 23 | Ad26.COV2.S | 28 days post dose | | 64% | | Moderate/ severe disease | | 81.7% | | Randomized controlled trial | Table 3 |
| 24 | BNT162b2 | 14 days post 2nd dose | 92%* | | 79%δ | PCR confirmed infection | Not reported | | | Test negative case control | Text |
| | ChAdOx1 nCoV-19 | 14 days post 2nd dose | 73%* | | 60%δ | | | | | | |

*S-gene "target failure" PCR used as indicator of B.1.1.7, # Assumed to be B.1.351 if not B.1.117, δ S-gene positive PCR assumed to be B.1.617.2
^ HIV-negative population

### Supplementary Table 3:

| Source of uncertainty | | Parameter in model | Estimated value | Standard error in value | Reference |
|---|---|---|---|---|---|
| Model parameters ($\log_{10}$ of the 50% protective neutralisation titre and natural log of the logistic slope parameter) | Symptomatic | $n_{50}, k_{log}$ | 1.13, -0.697 | Covariance matrix from fit | 25 |
| | Severe | $n_{50}, k_{log}$ | 1.12, -1.509 | Covariance matrix from fit | 25 |
| Standard deviation of neutralisation titres for each vaccine | | $\sigma_{all}$ | 0.46 | 0.022 | 25 |
| Mean ($\log_{10}$) neutralisation level of a vaccine against ancestral virus | | $\mu_s$ | Table S4 of reference (Ref) | Max. value across studies, 0.20 | 25 |
| Fold-change ($\log_{10}$) in neutralisation titre against each variant | | $\log_{10} \bar{F}^v$ | Fig. S3 | 0.015-0.029 | Current study |

Table S3: Sources of uncertainty in model predictions of vaccine efficacy against variants. In this table we use $k_{log}$ to denote $\log_e(k)$.

### Supplementary Table 4:

| Reference | Vaccine | Reference Group | Boosted Group (days post-infection) | Measured on day post-dose | Assay | Fold change (Boosted/ Reference) for ancestral virus | Neutralisation titre (fold convalescent) in reference group (Phase I/II trials)[25] | Predicted mean neutralisation level after boosting (fold convalescent) | Date derived from | Data on Variants |
|---|---|---|---|---|---|---|---|---|---|---|
| 26 | BNT162b2 | Naïve, 2 doses | Convalescent, 1 dose (mean:111) | 10 | Live virus Microneutralisation test, 50% endpoint titre | 4.8 | 2.4 | 11.4 | GMT titres in text | |
| 27 | BNT162b2 | Naïve, 2 doses | Convalescent, 2 doses (median: 53) | >14 | Live virus, Focus reduction neutralisation test (FRNT50) | 3.4 | 2.4 | 8.0 | Extracted from Figure 2 | Alpha, Beta, Gamma |
| 28 | BNT162b2 or mRNA-1273 | Naïve, 2 doses | Convalescent, 2 doses (median: 247) | 19/13 (reference/ convalescent) | Pseudovirus neutralisation | 5 | 3.1* | 15.7 | Mean value given in text | Beta |
| 29 | BNT162b2 or mRNA-1273 | Naïve, 2 doses | Convalescent, 2 doses (65-275) | 7 | Pseudovirus neutralisation | 9.2 | 3.1* | 28.7 | Raw data in supplement appendix | Beta |
| 30 | BNT162b2 or mRNA-1273 | Naïve, 2 doses | Convalescent, 2 doses | 28 | Live virus, Plaque reduction neutralisation assay (PRNT50) | 2 | 3.1* | 6.1 | Raw data kindly supplied | Alpha, Beta, Gamma, Delta, Epsilon, Eta, Iota, Kappa |
| 31 and 32 | mRNA-1273 (50 µg) | Naïve, 2 doses | mRNA-1273 boosted (177-226 post second dose) | 7/15 (reference/boost) | Pseudovirus neutralisation | 2.5 | 4.1 | 10.1 | Figure 3 (31) and Figure 1 (32) | Beta, Gamma |
| 33 | CoronaVac | Naïve, 2 doses | CoronaVac 3rd dose, (180 post-second dose) | 14 | Live virus, microneutralisation test | 4.9 | 0.17 | 0.82 | Figure 2 | |

* Geometric mean of neutralisation levels for BNT162b2 and mRNA-1273 reported in Phase I/II trials as summarised in reference 25
